# Considerations for healthcare wastewater surveillance of targeted antimicrobial-resistant organisms

**DOI:** 10.1101/2025.06.27.25330422

**Authors:** Angela Coulliette-Salmond, Florence Whitehill, Amanda K. Lyons, Bethelhem Abera, Colin Adler, Maroya Spalding Walters, Magdalena Medrzycki, Christine Ganim Kyros, Mariya Campbell, Michael Y. Lin, Rachel S. Poretsky, Adam Horton, Jennifer Weidhaas, James VanDerslice, L. Scott Benson, Erin M. Driver, Rolf U. Halden, Kerry A. Hamilton, Margaret Williams

## Abstract

Wastewater surveillance (WWS) at healthcare facilities paired with CDC-recommended patient screening methods may enable earlier detection of targeted antimicrobial resistant (AR) pathogens. However, this is a nascent field with knowledge gaps for how to best evaluate, implement, and interpret healthcare facility WWS. AR pathogens, such as carbapenemase-producing organisms and *C. auris*, are important non-viral targets in long-term acute care hospitals and ventilator-capable skilled nursing facilities (SNF) affecting high-risk, high-acuity patients and residents, respectively. To address the knowledge gaps, healthcare facility WWS was piloted at three Georgia SNFs (6 months per facility, weekly sampling), including obtaining permissions from health departments and facility administrators, conducting tracer studies to confirm effluent source, and evaluating sample collection approaches to manage low-flow and flushed healthcare debris. Additionally, a wastewater access survey was piloted in five states at 16 post-acute/long-term care facilities to assess feasibility of WWS regarding physical on-site manhole access, safety aspects, effluent flow, and other factors; 75% (12 of 16) of surveyed facilities demonstrated feasibility. Our findings suggest the importance of conducting thorough wastewater assay validation, assessing state public health lab capacity, as well as including paired wastewater samples with facility screenings in real-time to interpret the WWS results and understand the correlations with AR presence/prevalence among the facility population. Presented lessons learned and considerations are intended for stakeholders to review before implementing healthcare facility-level WWS for the purpose of AR organism public health surveillance to achieve the intended public health impact.

## Introduction

Data from 2017 showed the impact of carbapenem-resistant Enterobacteriaceae with 13,100 cases in hospitalized patients and an estimated $130 million in attributable healthcare costs^1^. There were 4,514 reported *Candida auris* clinical cases in 2023, which is concerning given that this fungal pathogen was first identified in 2013 ^2^. Facilities that care for high-acuity patients with long lengths of stay are considered ‘influential facilities’ with a disproportionate impact on regional transmission, where long lengths of stay are typically defined as >30 days ^3,4,5^. Models demonstrate that earlier detection with implementation of targeted infection control practices would reduce regional spread ^6,7,8,9,10^. Healthcare wastewater surveillance (WWS) is a notable early approach that has potential utility to support clinical screening practices and inform public health stakeholders on circulating and emerging antimicrobial resistant (AR) organisms, including *C. auris*. A few healthcare facility WWS studies for SARS-CoV-2 have approached the topic but have focused on the association between wastewater results and COVID-19 cases ^11,12,13,14^. However, there is limited information on elements to incorporate into healthcare WWS to achieve the intended impact. Thereby, the utility of WWS for the proactive detection of healthcare AR threats at the healthcare facility has come to the forefront ^15,16^.

Before initiating healthcare facility WWS, several engineering and field aspects should be considered. The facility building and plumbing, for example, likely did not consider WWS in design decisions and may not include a sewer manhole on the property. Historical knowledge and building blueprints are often unavailable prior to WWS implementation, thereby leaving the confirmation of the facility population captured by WWS uncertain. Additionally, wastewater sampling approaches have not been fully optimized at the healthcare facility level to address intermittent, low-flow, and healthcare-specific debris. Particularly for autosamplers, there are unforeseen complications at healthcare facilities due to the autosampler design and software program intended for a fully submerged vacuum line (Teledyne ISCO, personal communication), impeding the ability to collect a 24-h composite sample in low-flow settings. Combined with the nuances above, safety aspects of collecting wastewater and overall feasibility of conducting WWS at a healthcare facility are additional considerations.

Determining the utility of healthcare WWS is an important initial phase prior to utilizing these data to inform public health measures. It is critical to develop robust study designs that pair wastewater data with epidemiological and patient screening data from the healthcare facility, enabling accurate correlation analyses of wastewater signals with disease dynamics. Collecting these data is most easily accomplished after laying the groundwork for strong communication and collaboration between the WWS team, the facility, and the jurisdictional health department. Presented here are lessons learned and key considerations based on data collected in collaboration with 16 U.S. post-acute/long-term care facilities from 2022 to 2024 during healthcare WWS efforts for implementation specific to AR organisms of public health concern.

## Materials and Methods

This effort met the US Centers for Disease Control and Prevention (CDC) safety policies and recommendations in the CDC/NIH *Biosafety in Microbiological and Biomedical Laboratories* ^17^. This activity was reviewed by CDC, deemed research not involving human subjects, and was conducted consistent with applicable federal law and CDC policy^§^.

### Healthcare WWS partners

The Healthcare-Wastewater Antimicrobial Resistant Network (H-WARN) program within the Division of Healthcare Quality Promotion at CDC (Atlanta, GA) collaborated with the Georgia (GA) Department of Public Health (GDPH), Rush University Medical Center in partnership with the University of Illinois - Chicago, the University of Utah, and the Arizona (AZ) State University (ASU) to conduct WWS at post-acute/long-term care facilities. In this paper, the seven GA facilities were identified as Facility A to G, the five Illinois (IL) facilities were identified as Facility H to L, the Utah (UT) and Texas (TX) facilities were identified as Facility M and N, respectively, and the two AZ facilities were identified as Facility O and P. Types of care provided by each facility are described in Table 1 and individual state partnerships and facility selection are described in Supplement 1 (S1).

**Table 1.** Healthcare facility types and provided care for the best practices assessment (Facilities A, F, and G; n=3) and site surveys and feasibility determination (Facilities A to P; n=16), noting the following abbreviations: SNF - skilled nursing facility, ND - no data, LTACH - long-term acute care hospital, and RH - rehabilitation hospital.

| Facility ID | Facility type | Provided care |
| --- | --- | --- |
| A<sup>1,2</sup> | SNF | Skilled nursing, wound/ostomy, indwelling medical devices<sup>3</sup> |
| B, C | SNF | ND |
| D, F<sup>1</sup> | SNF | Skilled nursing, inpatient rehabilitation, wound/ostomy, indwelling medical devices<sup>3</sup> |
| E | vSNF | Skilled nursing, inpatient rehabilitation, wound/ostomy, ventilator care, indwelling medical devices<sup>3</sup> |
| G<sup>1</sup> | SNF | skilled nursing, inpatient rehabilitation, wound/ostomy, indwelling medical devices<sup>3</sup>, tracheostomy |
| H, I, J, K, L | LTACH | Long-term care, ventilator care |
| M | RH | Skilled nursing, inpatient and outpatient rehabilitation, wound/ostomy, indwelling medical devices<sup>3</sup> |
| N | vSNF devices<sup>2</sup> | Skilled nursing, wound/ostomy, indwelling medical |
| O | Mixed-use | Skilled nursing, memory care, assisted living, and independent living units |
| P | Hospital | Emergency care, critical care, advanced surgical procedures, pain management, wound care |

<sup>1</sup> facilities participating in the tracer study; <sup>2</sup> facility participating in low-flow strainer assessment; <sup>3</sup> indwelling medical devices: central vascular catheter, urinary catheter, etc.

### Lessons learned – Tracer studies and strainer assessment

*Facility descriptions:* Three skilled nursing facilities (A, F, and G) in the Atlanta metro area (GA) participated in the tracer studies, while the strainer assessment was conducted only at Facility A.

*Tracer studies:* The two-part tracer study included a fluorescent tracer dye (Bright Dyes^®^ FLT yellow/green tablet; Kingscote Chemicals, Miamisburg, OH) followed by a biological surrogate heat-inactivated (60±2°C for 40 min) Inforce^®^ 3 Respiratory lyophilized calf vaccine (Parsippany, New Jersey; Zoetis) on a subsequent day. The tracer study was used to gauge wastewater residence time (i.e., amount of time wastewater spends in pipes between toilet and collection/sampling points) after toilet flush and confirm that the selected manholes captured the intended population at Facilities A, F, and G. Methods are further described in S1.

*Strainer assessment*: Three strainers compatible for use with the Teledyne ISCO Avalanche Autosampler (Lincoln, NE) were evaluated at Facility A with the aim of consistent collection of the desired wastewater sample volume (Figure 1). The Standard Weighted Polypropylene Strainer (“Standard”, n=9 volume comparisons between ‘expected’ and ‘collected’; Lincoln, NE; Teledyne ISCO), Low Flow Stainless Steel Strainer (“Low Flow”, n=7; Lincoln, NE; Teledyne ISCO), and a custom fabricated strainer (“Custom”, n=9; Atlanta, GA; CDC) were deployed, and ‘expected’ and ‘collected’ sample volumes were recorded. See Figure 1 for strainer visuals and design differences; the Custom strainer technical drawing in Supplemental 2 (S2); and methods and statistics are further described in S1.

**Figure 1.**
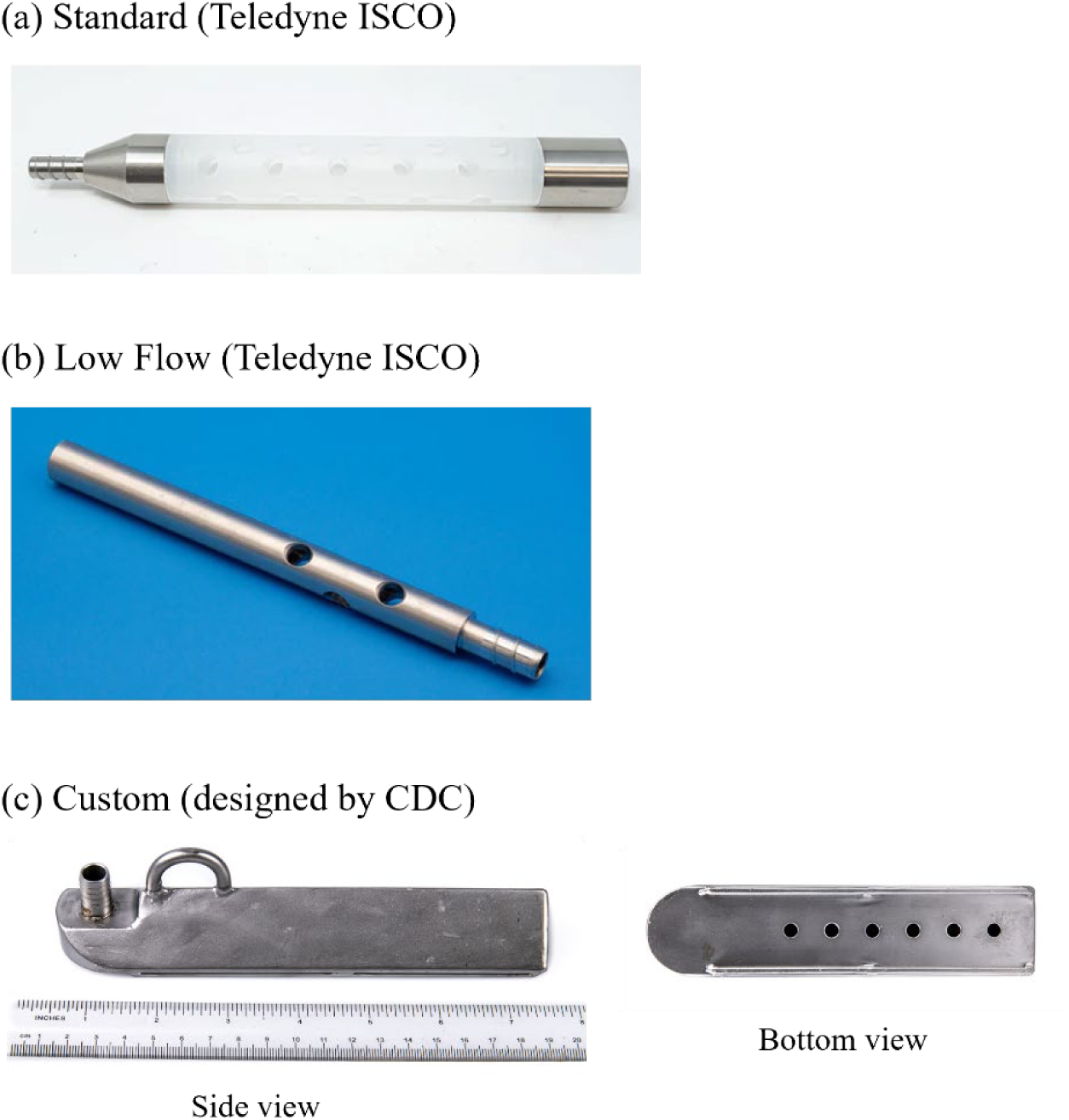
Strainer types, (a) Standard (photo: https://store.teledyneisco.com) – standard weighted polypropylene strainer (3/8” ID); (b) Low Flow (photo: https://store.teledyneisco.com) – low flow stainless steel strainer (3/8” ID); and (c) CDC custom

### WWS feasibility assessment

*Pilot survey*: In 2021, H-WARN developed an 8-question pilot survey. This was administered onsite at seven GA post-acute care facilities (Facilities A-G) between 2021 and 2022, where the primary respondents included facility administrators, directors, nurses, and infection control specialists (Table 1). The survey evaluated potential sampling locations conditions, safety concerns for sampling wastewater at these locations, facility information regarding types of care, MDRO screening practices, daily visitor counts, and fecal-waste management practices.

*Expanded pilot survey*: Starting in March 2023, partners in IL and UT collaborated with H-WARN to incorporate additional data elements and develop a new set of questions (e.g., similar data collection aspects using different questions). The 28-question expanded pilot survey was administered during in-person site visits at 9 post-acute care facilities in IL (5), UT (1), TX (1), and AZ (2) that included LTACHs, SNFs, ventilator capable SNFs (vSNFs), as well as a rehabilitation hospital (RH), mixed use facility, and general hospital. The survey new sections covered administrative elements, willingness to participate, wastewater access, grease traps/drainage lines, and plumbing aspects. The expanded pilot survey questions are provided in the Supplemental 3 (S3).

*Feasibility*: Feasibility was determined by three main factors: facility-level wastewater access availability, wastewater access practicality, and facility administration amenability.

Wastewater access availability incorporated understanding if a sewer manhole was on facility property, whether the access point wastewater captured the target population, and if the condition of the wastewater access point and piping were compatible for safe collection and use of sampling equipment. Wastewater access practicality considered whether there was ample room for sampling personnel and the required equipment, the location was safe for sampling with respect to the sampling personnel, the facility residents/patients, visitors, and healthcare personnel, and that the regular function of the facility was not interrupted. The overall facility amenability, meaning a willingness to participate at both facility-level and corporate levels for the duration of the planned WWS, includes considering consistent point(s) of contact (POC(s)), weekly questionnaires completion, and communication to explain the study purpose for the impacted audience (i.e., residents, visitors, staff).

### Key considerations

The key considerations were developed through the WWS efforts presented here and collaborations with healthcare facilities, funded partners, health departments, and others who have subject matter expertise regarding antimicrobial resistant organisms, clinical screening, WWS, laboratory networks, and healthcare systems.

## Results

### Lessons learned – Tracer studies and strainer assessment

The tracer study was conducted at Facility A, F, and G, while the low-flow assessment was conducted at Facility A only. Facility A and Facility F were free-standing, multilevel buildings, and Facility G was a free-standing building with one level.

*Tracer studies:* The studies were conducted in toilets on the fifth floor (topmost floor housing residents) of Facility A (n=2 tracer studies) and the ground floor (only floor housing residents) of Facility G (n=1). At Facility F (n=3), the first dye tracer study was unsuccessful on first and fourth floor bathrooms from the middle of the facility and not visually observed at the manhole that the facility believed would capture the intended section of the building. As a result, a thorough walk-through of facility plumbing details, and evaluation of facility maps and assessment of manhole locations were conducted. The subsequent tracer study at Facility F involved two ground-floor toilets in separate facility wings, with simultaneous observation at two manholes. This effort was successful and confirmed the proper manholes for WWS in Facility F. Overall, the three facilities had visual detection between 1.5 and 2.5 minutes from toilet flush and dissipation of the dye tracer between 4 and 7 minutes from initial visual detection (Table 2).

**Table 2.** Tracer studies conducted at Georgia skilled nursing facilities, where the visual dye using the fluorescent tracer dye (Bright Dyes^®^ FLT yellow/green tablet; Kingscote Chemicals, Miamisburg, OH) and viral tracer studies using heat-inactivated Inforce^®^ 3 Respiratory Vaccine lyophilized calf vaccine (Parsippany, New Jersey; Zoetis) with bovine respiratory syncytial virus (BRSV) as the detected target were conducted at Facility A (5 floors, n=17 samples), Facility F (4 floors, n=16 samples), and (iii) Facility G (1 floor, n=19 samples). Noted are the floor the tracers were introduced into the toilets, detection and peak detection (min), as well as the dissipation time (min).

|  | Visual dye tracer |  |  | Viral tracer (n=3 per facility) |  |
| --- | --- | --- | --- | --- | --- |
|  | n | Detection | Dissipation | Peak detection | Dissipation |
| Facility A<sup>1</sup><br>5<sup>th</sup> floor | 2 | 2.5 min | 7 min | 2 to 10 min<br>(1.35 x 10<sup>3</sup> to 1.13 x 10<sup>2</sup> gc/ µl) | 40 min |
| Facility F<sup>2</sup><br>1<sup>st</sup> floor | 3 | 2.0 min | 5.5 min | 2 to 15 min<br>(2.58 x 12<sup>2</sup> to 1.57 x 10<sup>2</sup> gc/ µl) | 90 min |
| Facility G<sup>3</sup><br>1<sup>st</sup> floor | 1 | 1.5 min | 4.0 min | 2 to 10 min<br>(6.93 x 10<sup>4</sup> to 4.18 x 10<sup>2</sup> gc/ µl) | 240 min |

<sup>1</sup> Facility A sampling time points: 0, 1, 2, 3, 4, 5, 6, 8, 10, 15, 30, 35, 40, 45, 60, 90, 120 minutes
<sup>2</sup> Facility F sampling timepoints: 0, 1, 2, 3, 4, 5, 6, 8, 10, 12, 15, 30, 45, 60, 90, 120 minutes
<sup>3</sup> Facility G sampling time points: 0, 1, 2, 3, 4, 5, 6, 8, 10, 30, 45, 60, 120, 150, 180, 240, 720, 1080, 1440 minutes

The biological surrogate tracer studies were subsequently performed at Facility A, F, and G (n=3 studies per facility) in the same successful locations as the visual dye tracer (Table 2).

Facility A (n=17 samples) and F (n=16 samples) studies were conducted over a 120-min time window, while Facility G (n=19 samples) was performed over a 1440 min (24 h) time window. Initial peak biological surrogate concentrations were detected at 2 min at all facilities with a decreased peak between 10-15 min and dissipation time points ranging from 40 min at Facility A to 240 min at Facility G. At Facility G after the initial decline, there were notable concentration increases between 150 and 180 minutes (2.23 x 10^2^ - 4.95 x 10^2^ gc/ µl) before continuing to decline and the reason for the extension to 1440 minutes.

*Strainer assessment:* The Standard strainer was deployed for three weeks (n=9 volume comparisons between ‘expected’ and ‘collected’), the Low-flow strainer for two weeks (n=7), and the Custom strainer for three weeks (n= 9). Complications with the Standard and Low-flow strainers included the strainer collection holes being blocked by healthcare wipes and the collection holes not being fully immersed, thereby inhibiting wastewater collection and often creating air bubbles in the line, which caused inaccurate collection. The Custom strainer, while covered in healthcare wipes, was capable of wastewater collection due to the design with collection holes exclusively on the bottom (i.e., fully immersed in the low flow wastewater) with mini-rails lifting the strainer off the bottom sewer by millimeters, allowing consistent and accurate sample collection (Fig. 2). These design differences were reflected in the expected (programmed) versus collected volumes (L), where the volume differences for Standard, Low-flow, and Custom strainers were -0.18 L (±0.61), -6.18 L (±4.14), and -0.04 L (±0.36), respectively. Significant differences calculated using the absolute values of the mean volume differences were demonstrated between all three strainer types using the chi-square test of independence (χ^2^(2)=17.8; p<0.05): Standard and Low-flow (p-value<0.05), Standard and Custom (p-value<0.05), and Low-Flow and Custom (p-value<0.0001) strainers.

**Figure 2.**
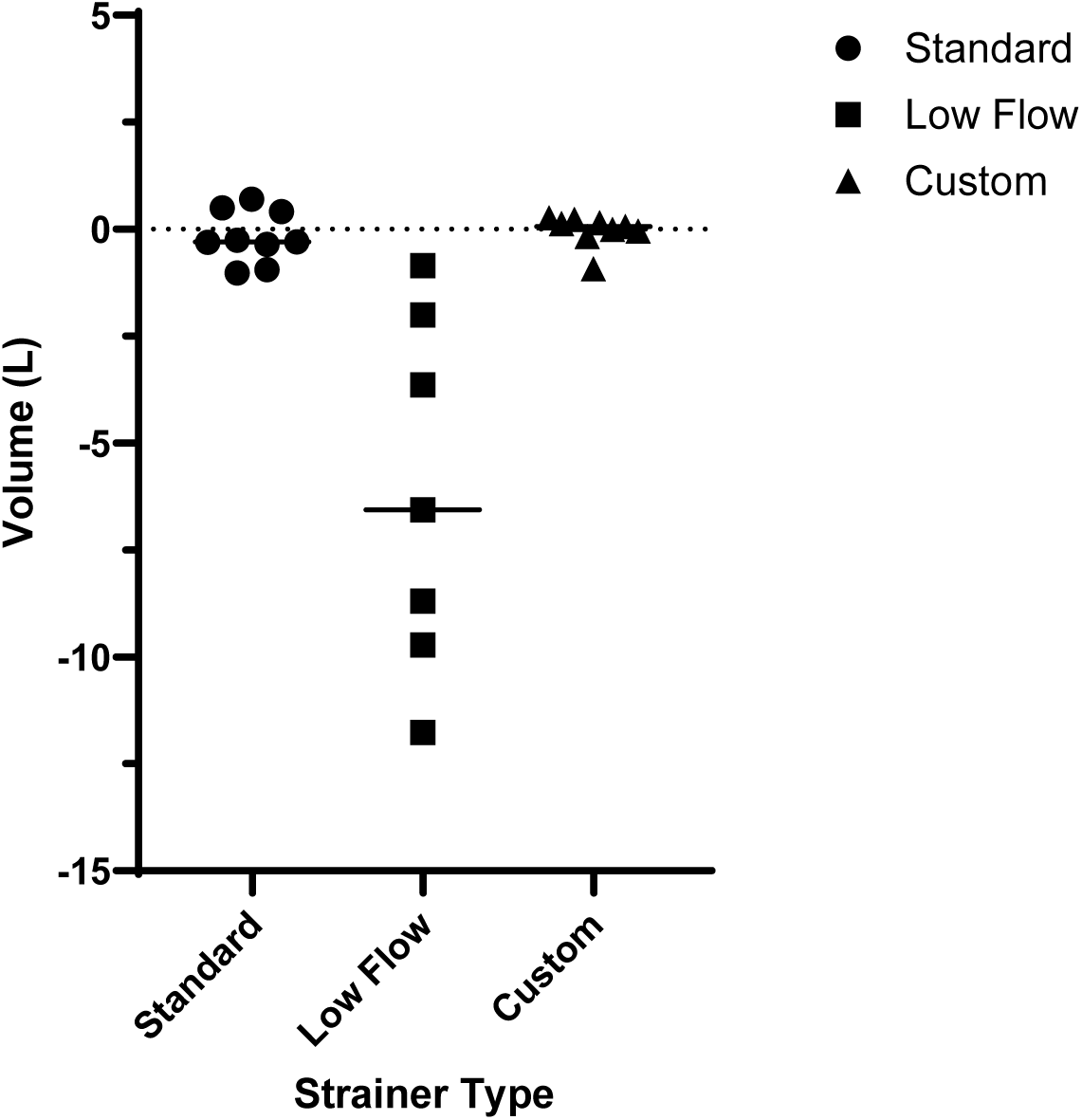
Strainer comparison of the “Standard” (n=9), “Low Flow” (n=7) and “Custom” (n=9) auto sampler compatible strainers for wastewater sampling, where the mean volume differences (expected versus collected) were -0.18 (±0.61), -6.18 (±4. 14), and -0.04 (±0.36) L, respectively.

### Feasibility assessment

The facility general descriptions, average census, facility types, and survey results are detailed in Tables 3 and 4, where footnotes provide specifics regarding access points, safety concerns, and extenuating circumstances that would make WWS not feasible. Results per state for the pilot and expanded surveys are detailed in S1.

**Table 3.** Pilot survey results at seven Georgia (USA) post-acute care facilities to understand potential sampling locations conditions and any safety concerns for sampling wastewater.

| Facility | A | B | C | D | E | F | G |
| --- | --- | --- | --- | --- | --- | --- | --- |
| **Resident Census N** | 169 | 107 | 99 | 61 | 109 | 118 | 92 |
| **Number of Residents Contributing Fecal Material to Wastewater n (%)** | 66 (40%) | 16 (15%) | ND | 20 (33%) | 21 (19%) | Approx 85% | Approx 60–70% |
| **Manhole Appropriate for Sampling?** | Yes | Yes<sup>1</sup> | Yes<sup>1,2</sup> | Yes | Yes<sup>1</sup> | Yes | Yes |
| **Wastewater Flow Adequate for Sampling?** | Yes | Yes | No<sup>3</sup> | Yes | Yes | Yes | Yes |
| **Cleanout Accessible for Sampling?** | Yes | No<sup>4</sup> | No<sup>4</sup> | Yes | No<sup>4</sup> | No<sup>5</sup> | ND |
| **Facility Amenable to Project?** | Yes | Yes | Yes | ND<sup>6</sup> | Yes | Yes | Yes |
| **Open Communication Avenues at Facility?** | Yes | Yes | Yes | ND<sup>6</sup> | Yes | Yes | Yes |
| **Safe Sampling Site?** | Yes | Yes | No<sup>7</sup> | No<sup>8</sup> | No<sup>9</sup> | Yes | Yes |
| **Overall, is sampling wastewater feasible at this facility?** | **Yes** | **Yes** | **No** | **No** | **No** | **Yes** | **Yes** |

**Abbreviations:** ND = not determined;
<sup>1</sup>No nearby electrical outlet for autosampler.
<sup>2</sup>Large heavy square manhole covering was difficult to open with standard manhole opening tools.
<sup>3</sup>Low wastewater flow in manhole. A sludge mat several inches thick was covering the flow and would require a plumbing intervention to remove debris.
<sup>4</sup>No cleanout available or unable to locate cleanout.
<sup>5</sup>Cleanout located in the middle of the dining room.
<sup>6</sup>Not assessed because sampling site was unsafe and therefore sampling could not be performed at this facility.
<sup>7</sup>Out of the way of pedestrian and vehicular traffic but the non-standard square manhole covering could pose a safety risk.
<sup>8</sup>Both manholes on the property were in unsafe locations. One was near the entrance of the facility and the other was close to a street with vehicular traffic.
<sup>9</sup>Effluent pipe was broken about two feet below the manhole opening which created a safety concern for wastewater splashing out of manhole.

**Table 4.** An excerpt of expanded pilot survey results at a total of nine post-acute care facilities in Illinois (IL), Utah (UT), Texas (TX), and Arizona (AZ) (USA) focusing on the feasibility for wastewater sampling. See the Supplemental 3 for the entire expanded survey questions.

| Facility | H | I | J | K | L | M | N | O | P |
| --- | --- | --- | --- | --- | --- | --- | --- | --- | --- |
| State | IL | IL | IL | IL | IL | UT | TX | AZ | AZ |
| (Q1) Resident census | 59 | 98 | 80 | 28 | 30 | 45 | 85 | 11 | 120 |
| (Q5) Amenable to WW equipment? | Yes | Yes | Yes | Yes | Yes | Yes | Yes | Yes | Yes |
| (Q6) Amenable to informational signs and fliers? | Yes | Yes | Yes | Yes | Yes | Yes | Yes | Yes | Yes |
| (Q13) Is there at least one external physical access point to the WW? | Yes<sup>1</sup> | Yes<sup>2</sup> | No | Yes<sup>1</sup> | Yes<sup>1</sup> | Yes<sup>1</sup> | Yes<sup>3</sup> | Yes | Yes |
| (Q14) Where is the external WW access point located? | On land-scaped grounds | On land-scaped grounds | n/a<sup>4</sup> | Public sidewalk | On land-scaped grounds | By dumpsters at loading dock, away from traffic | CO at end of East wing; MH in roadway of parking lot in back of facility | On land-scaped grounds | In parking lot, but where there is traffic |
| (Q15) Approximately how far from the building is the WW access point? | 20–50 ft | <20 ft | n/a<sup>4</sup> | <20 ft | >100 ft | 20 ft | CO 6 ft; MH 40 ft | 20–50 ft | <20 ft |
| (Q16) Manhole cover |  |  |  |  |  |  |  |  |  |
| a. Energy source/outlet? | No | Yes | n/a<sup>4</sup> | No | No | No | No | No | No |
| b. Description of manhole |  |  |  |  |  |  |  |  |  |
| MH cover diameter | 22.75 in | 22.75 in | n/a<sup>4</sup> | 22.75 in | 22.75 in | 22.75 in | 24 in | 30 in | 24 in |
| Depth | 13 ft | 20 ft | n/a<sup>4</sup> | 4 ft | 13 ft | 11 ft | 11 ft | 8 ft | 6 ft |
| (Q17) Sufficient space for setting up equipment? |  |  |  |  |  |  |  |  |  |
| a. Autosampler | Yes | Yes | n/a<sup>4</sup> | Yes | Yes | Yes<sup>5</sup> | Yes<sup>5,6</sup> | Yes | Yes |
| b. Passive | Yes | Yes | n/a<sup>4</sup> | Yes | Yes | Yes<sup>5</sup> | Yes<sup>5,6</sup> | Yes | Yes |
| (Q18) Any feasibility concerns? | No | No | Yes<sup>4</sup> | Yes<sup>7</sup> | No | Yes<sup>8</sup> | Yes<sup>9</sup> | Pedestrian traffic<sup>10</sup> commingled flow<sup>11</sup> | Parking lot and daily traffic |
| Overall, is sampling wastewater feasible at this facility? | Yes | Yes | No | Yes | Yes | Yes | Yes | Yes | Yes |

**Abbreviations:** WW = wastewater, CO = cleanout, MH = manhole, n/a = not applicable;
<sup>1</sup>Access point is a manhole.
<sup>2</sup>Access point is a manhole lift station.
<sup>3</sup>One manhole and one cleanout.
<sup>4</sup>No external physical access point at this facility.
<sup>5</sup>There is room for sampling equipment if it is placed in the manhole.
<sup>6</sup>There is room for sampling equipment next to the building near the cleanout.
<sup>7</sup>Location is in the middle of a public sidewalk.
<sup>8</sup>Sampling equipment could obstruct garbage pickup location.
<sup>9</sup>Manhole is in parking lot therefore would need a vehicular traffic diversion. Area could flood with heavy rain.
<sup>10</sup>Barricades must be used at all times due to pedestrian traffic.
<sup>11</sup>Two wastewater drainage pipes merge and concerted effort must occur to make sure passive and active sampling is within the comingled flow.

*Overall Feasibility (Combined Facilities A - P)*: Twelve (75%) of 16 total facilities surveyed (pilot and expanded pilot surveys) determined that WWS was feasible. Of the 16 total healthcare facilities surveyed, 15 (93.8%) had at least one external physical access point with wastewater access availability, where 10 (66.7%) of the 15 were located on landscape grounds versus a paved area with pedestrian or vehicle traffic. Focusing on the nine facilities included in the expanded pilot survey, only one facility (11%) had a nearby outlet available to power an autosampler. The most common safety concerns centered on the wastewater access being a manhole located in an area with pedestrian or vehicle traffic, and manhole covers being of atypical size/shapes that would interfere with typical wastewater sampling equipment. Of the eight expanded pilot surveyed facilities with wastewater access availability (Facility J did not have an onsite wastewater access point), five (62.5%) had laundry services (two sites had bleach injectors and one had a dedicated bedside commode disinfection room). Facility amenability was noted for all 9 (100%) expanded pilot surveyed facilities. Of note, those conducting WWS (e.g., academia) without established links to public health (i.e., post-acute/long-term care facilities) had more challenges with engagement than the partners who had existing partnerships.

### Key considerations: WWS for antimicrobial resistant organisms

The aforementioned learned lessons and feasibility assessment described are pathogen agnostic and applicable for generic WWS at post-acute/long-term care facilities. However, key considerations to determine the utility of WWS at healthcare facilities for antimicrobial resistant organisms that are commonly healthcare associated are outlined in Box 1. The goal of correlating AR prevalence from WWS data to the prevalence of AR from patient screening or clinical culture data would be to allow WWS to reduce the need for data from other sources.

A vital primary step, combined with the knowledge gained from a tracer study and a feasibility assessment, should emphasize alignment of wastewater with epidemiological and correlation components. It is critical that facility-level epidemiological metrics are collected to understand the dynamics of the population contributing to the wastewater signal. These epidemiological data include, but are not limited to, current prevalence of targeted AR organisms (e.g., carbapenemase-producing organism, *C. auris*), the number of patients/residents who use the toilet and bathe/shower, and the waste management practices (i.e., facility methods for disposing of fecal wastes from bed pans, briefs, and bed baths). Communication includes consistent messaging in emails to recruit facilities, provide informational study flyers, signage on field equipment, and monthly reports to the facility. An additional consideration for communication materials is to have one designated POC at the facility to manage WWS-related communications. It is recommended that stakeholders (e.g., LTCF, wastewater experts, HDs) take the time to draft a plan that outlines a course of action for each type of WWS result, noting that response plans be specific to the target. Public health has an important role in understanding and responding to AR and that when designing WSS at healthcare facilities, consulting with local and state authorities is recommended due to their knowledge and trusted relationships with the LTACHs/SNFs, facilities at risk for the target organism, and where identifying the target organisms may inform prevention and response activities. Once all stakeholders have committed to engagement in a healthcare WWS effort, written agreements with defined roles and responsibilities are encouraged to establish a clear understanding of expectations, workflows, timelines, data management, results sharing, and response plan.

Alignment of wastewater samples and point prevalence surveys (PPSs; i.e., rectal swab screening of a ward or facility using verbal consent) from paired same-day collection to result analysis are critical to build evidence towards the utility determination of WWS at healthcare facilities for desired targets. Of note, it is important to utilize a tracer study confirmed onsite wastewater collection site that verifies the unit where the PPS is being performed. Stakeholders should decide which approach works most effectively depending on the timing of the PPS and whether screening will be performed across multiple days (e.g., due to large facility size or staffing constraints): wastewater sampling (a) within a 24- to 48-h window of the PPS or (b) the days of PPS start and completion. Another consideration is understanding how the facility ascertains prevalence and their approaches to assess transmission in an outbreak setting, to allow for coordination with the wastewater group to conduct additional sampling and/or share aggregate clinical data. A higher frequency of wastewater sampling, in addition to the paired sampling (wastewater and PPSs), is also proposed to adequately track wastewater background signal. As mentioned, this is a nascent field and assessing when a correlation determination is finalized per AR target, facility type, geographic region, etc. will likely be an ongoing conversation among stakeholders.

Assays used for target detection to evaluate the alignment between wastewater and PPSs and/or patient screenings are also important. Open discussions should occur between the wastewater and public health labs regarding laboratory assays used and coordination for the wastewater laboratory to evaluate detection assays using the wastewater matrix for testing (i.e., concentrate, extract, and/or lysates). Full characterization and validation of assays with positive controls in wastewater matrices using consistent performance criteria and optimization processes to understand variation in laboratory performance, limit of detection, limit of quantification, recovery efficiency across sites, etc., will improve antimicrobial resistant organism assays for WWS. Ideally, inter- and intra-laboratory comparisons using the same primer/probe sequences and detection platforms would be performed for confidence in the wastewater results to allow more fluent analysis in determining the utility of healthcare WWS.

## Conclusions

The National Academies report acknowledges that facility-level AR WWS would likely be more actionable particularly for antimicrobial resistant organisms for vulnerable populations than community-level WWS but this has not been robustly tested ^16^. A main goal with the information presented is to assure that as other programs initiate WWS at healthcare facilities, there are foundational considerations to reference. The tracer study revealed inaccurate sewer plumbing knowledge for one of three facilities, emphasizing the importance of confirming a wastewater effluent source before initiating WWS efforts that will be associated with clinical data. This ‘lesson learned’ was relayed to the Illinois, Utah, and Arizona WWS partners, who successfully conducted visual dye tracer studies to confirm wastewater collection sites. The low-flow strainer development provided accurate, consistent wastewater collection for a 24-hour composite sample using an autosampler at a healthcare facility. This highlights that wastewater collection approaches that have been designed for environmental waters and community-level WWS do not always translate to usability at the facility-level. Passive sampling approach may provide an alternative to low-flow issues but may not be feasible if exact flow conditions and volumes cannot be determined. Additionally, in association with conducting a tracer study and application of appropriate strainer for sampling, conducting an onsite, in-person survey to understand the feasibility is vital to incorporate the array of field aspects, safety considerations, and facility willingness. The 75% feasibility determination from 16 facilities in five states provides a starting point for assessing the potential of broader WWS at healthcare facilities for increased surveillance of emerging antimicrobial resistant organism surveillance to support clinical screenings and future response capabilities. However, the survey needs to be administered more widely and systematically to have an increased resolution of national capacity.

For the utility determination and potential future use of healthcare WWS, these key considerations bolster analysis of the correlation between wastewater data and clinical screening data. Additionally, if programs include these key considerations in a standardized approach, then comparing across programs will be feasible and enhance the ability to determine successful WWS approaches at healthcare facilities. The existence of integrated, public wastewater working groups that include academic, government, and private sectors (e.g., APHL Community of Practices, WEF wastewater summits) provide an avenue to unify healthcare WWS efforts. While challenges remain, such as addressing the confounding factor of microbial growth and biofilms in the sewer plumbing, broadening use of national feasibility surveys for response, and supporting community outreach to healthcare facilities with hesitancy to participate, WWS continues to hold promise for improving knowledge regarding our population’s public health.

## Data Availability

All data produced in the present study are available upon reasonable request to the authors

## Acknowledgements

First and foremost, the participating healthcare facilities in Georgia, Illinois, Utah, Texas, and Arizona deserve much appreciation for collaborating continuously through the WWS efforts. Additionally in Georgia, we extend gratitude to Jeanne Negley and Joanna Wagner at the Georgia Department of Public Health for their support and guidance during the initial phases of WWS during the COVID-19 pandemic. The credit in the creative design for of the CDC custom strainer goes to D. Neal Whaley Jr., in conjunction with Christine Ganim Kyros, and technical drawing designed by D. Neal Whaley, Jr. and drawn by Harris Sheinman. For those in the Division on Healthcare Quality Promotion (CDC, Atlanta, GA) who supported this effort, we thank you for your time and diligence during the COVID-19 pandemic. The Safety and Healthcare Epidemiology Prevention Research Development (SHEPheRD) contract managed by the Division of Healthcare Quality and Promotion Part funded a portion of this WWs effort to the Rush University Medical Center-Chicago, University of Utah, and Oak Ridge Associated Universities partners.

## Author contributions

ACS: Conceived of the presented idea, supervised lesson learned, conducted tracer study, designed pilot survey, contributed to expanded pilot survey, wrote the manuscript with support from FW, AL, BA, CA, MW, MM, CGK, MC, ML, RP, AH, JW, JVS, SB, ED, RH, KH, MW. FW, AL: analyzed the pilot and expanded pilot survey data; FW: designed considerations/key points; AL: conducted tracer study; BA, CA: data analysis of Georgia facility data and figure creation; MW, MM: contributed to considerations/key points; CGK: conducted tracer study and low-flow assessment; MC: conducted the tracer study, process laboratory samples; ML, RP, AH: conducted expanded pilot survey in Illinois and contributed to the feasibility assessment; JW, JVS, SB: conducted expanded pilot survey in Utah and Texas and contributed to the feasibility assessment; ED, RH, KH: conducted expanded pilot survey in Arizona and contributed to the feasibility assessment; MW: Contributed to and reviewed manuscript and associated poster/presentations.

## Footnote page

§See e.g., 45 C.F.R. part 46, 21 C.F.R. part 56; 42 U.S.C. §241(d); 5 U.S.C. §552a; 44 U.S.C.

§3501 et seq.

## Funding statement

This project was supported in part by an appointment to the Research Participation Program at the Centers for Disease Control and Prevention administered by the Oak Ridge Institute for Science and Education through an interagency agreement between the U.S. Department of Energy and the Centers for Disease Control and Prevention.

A portion of this study was funded by the CDC grant No. 75D30121D12772 to the Rush University Medical Center (ML), No. 75D30121D12774 to the University of Utah (JW), and No. 75D30121D12769 to the Oak Ridge Associated Universities, Inc. (RH). The funder had no role in conducting the study; collection, management, analysis of the data.

Meetings where the information has previously been presented

(Poster) Whitehill, F., A. Coulliette-Salmond, Amanda Lyons, Bethelhem Abera, Colin Adler, Judith Noble-Wang, Michael Lin, Rachel Poretsky, Adam Horton, James VanDerslice,

Jennifer Weidhaas, and Margaret Williams. “In-person wastewater access review survey at post-acute care facilities across three U.S. regions.” Water Environment Federation, Wastewater Disease Surveillance Summit. August 12-13, 2024. Atlanta, Ga.

(Presentation) Coulliette-Salmond, A. Overview presentation for Panel: “Wastewater surveillance at long-term care facilities: Perspectives on feasibility and antimicrobial resistant target detection.” American Public Health Laboratories (APHL) conference 2024. May 6-9, 2024. Milwaukee, Wisconsin.

(Poster) Coulliette, A.D., M. Burgos-Garay, Y. Cahela, M. Campbell, R. Donlan, C. Ganim Kyros, L. Franco, A. Keaton, S. Lenz, A. Lyons, M. Medrzycki, J. Moore, M. Walters, D. Neal Whaley, and J. Noble-Wang. “Assessment of challenges and implementation of best practices for skilled nursing facility wastewater surveillance.” Microbes in Wastewater: Molecular Approaches in Pathogen and AMR Surveillance. January 18-19, 2024. Laguna Beach, CA.

(Presentation) Georgia Emerging Infections Program. “Facility-Level Wastewater Surveillance: Strategies for Emerging Pathogens and Antimicrobial Resistance Genes/Organisms in Healthcare Settings.” Atlanta, GA. August 18, 2023.

---

**Box 1.**
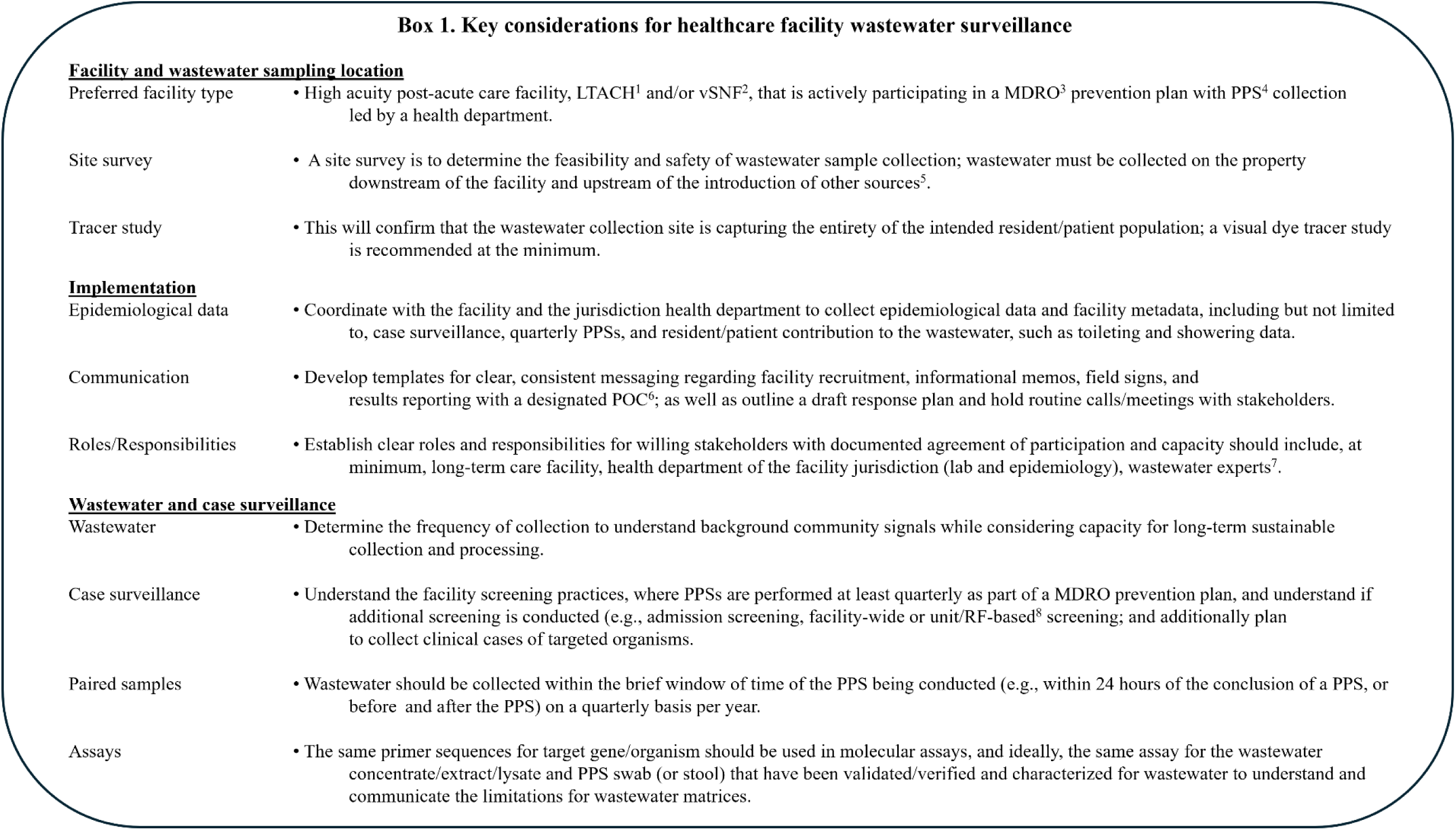
Key considerations for healthcare facility wastewater surveillance ^1^ LTACH: long-term acute care hospital; ^2^ ventilator capable skilled nursing facility; ^3^ MDRO: multidrug-resistant organism, ^4^ PPS: point prevalence survey, colonization screening performed unit- or facility-wide following the identification of a patient/resident with a novel or targeted MRDO, or for purposes of prevention; ^5^ site survey in Supplemental 2; ^6^ POC: point-of-contact; ^7^ wastewater experts defined: involvement in an active or previous WWS program, existing capacity to collect WW, laboratory capacity using appropriately validated concentration, extraction, and detection for designated AR targets, authors on a peer-reviewed publication, dashboard, or participation in the National Wastewater Surveillance System or similar programmatic efforts; ^8^ RF-based: risk factor-based

|  |  |
| --- | --- |
| Preferred facility type | • High acuity post-acute care facility, LTACH<sup>1</sup> and/or vSNF<sup>2</sup>, that is actively participating in a MDRO<sup>3</sup> prevention plan with PPS<sup>4</sup> collection led by a health department. |
| Site survey | • A site survey is to determine the feasibility and safety of wastewater sample collection; wastewater must be collected on the property downstream of the facility and upstream of the introduction of other sources<sup>5</sup>. |
| Tracer study | • This will confirm that the wastewater collection site is capturing the entirety of the intended resident/patient population; a visual dye tracer study is recommended at the minimum. |
| Epidemiological data | • Coordinate with the facility and the jurisdiction health department to collect epidemiological data and facility metadata, including but not limited to, case surveillance, quarterly PPSs, and resident/patient contribution to the wastewater, such as toileting and showering data. |
| Communication | • Develop templates for clear, consistent messaging regarding facility recruitment, informational memos, field signs, and results reporting with a designated POC<sup>6</sup>; as well as outline a draft response plan and hold routine calls/meetings with stakeholders. |
| Roles/Responsibilities | • Establish clear roles and responsibilities for willing stakeholders with documented agreement of participation and capacity should include, at minimum, long-term care facility, health department of the facility jurisdiction (lab and epidemiology), wastewater experts<sup>7</sup>. |
| Wastewater | • Determine the frequency of collection to understand background community signals while considering capacity for long-term sustainable collection and processing. |
| Case surveillance | • Understand the facility screening practices, where PPSs are performed at least quarterly as part of a MDRO prevention plan, and understand if additional screening is conducted (e.g., admission screening, facility-wide or unit/RF-based<sup>8</sup> screening; and additionally plan to collect clinical cases of targeted organisms. |
| Paired samples | • Wastewater should be collected within the brief window of time of the PPS being conducted (e.g., within 24 hours of the conclusion of a PPS, or before and after the PPS) on a quarterly basis per year. |
| Assays | • The same primer sequences for target gene/organism should be used in molecular assays, and ideally, the same assay for the wastewater concentrate/extract/lysate and PPS swab (or stool) that have been validated/verified and characterized for wastewater to understand and communicate the limitations for wastewater matrices. |

