## Supplemental for "Considerations for healthcare wastewater surveillance of targeted antimicrobial-resistant organisms"

**Supplemental 1.**

A. **Methods**
   State partnerships and facility selection
   Tracer studies
   Strainer assessment
B. **Feasibility individual state results**
   Pilot survey: Georgia
   Expanded survey: Illinois, Utah, Texas, Arizona

#### A. Methods

<u>State partnerships and facility selection</u>

The Healthcare-Wastewater Antimicrobial Resistant Network (H-WARN) program within the Division of Healthcare Quality Promotion at the CDC (Atlanta, GA) collaborated with the Georgia (GA) Department of Public Health (GDPH). The GDPH provided recommendations and initial communications with healthcare facilities via email. Once positive interest was indicated by healthcare facilities, H-WARN then led communications regarding the WWS effort to recruit for participation over virtual meetings and subsequent in-person site visits. The GA facilities were identified as Facility A to G.

WWS efforts in Illinois (IL) were led by Rush University Medical Center in partnership with University of Illinois Chicago and local public health departments (Chicago DPH and Cook County DPH). A convenience sample of LTACHs was identified through contacts known through prior and existing quality improvement collaborations. The IL facilities were identified as Facility H to L.

WWS efforts in Utah (UT) and Texas (TX) were led by the University of Utah. Contacts from the state health department and lab, individuals involved with statewide HAI efforts and contacts working for target facilities provided recommendations. These facilities were contacted, and initial and follow-up meetings were conducted in person or remotely. Several meetings were required to convince facilities to participate. The UT and TX facilities were identified as Facility M and N, respectively.

WWS efforts in Arizona were facilitated through a combined effort by Arizona State University (ASU); LeadingAge Arizona, a non-profit that advocates on issues of housing, healthcare, and home services for people 65+ years of age; and the City of Tempe Office of Strategic Management and Innovation that oversees wastewater surveillance activities within the community. The AZ facilities were identified as Facility O and P.

Tracer studies

A two-part tracer study, consisted of a visual dye tracer followed by a biological surrogate on a subsequent day, was used to gauge wastewater residence time in pipes after toilet flush and confirm the selected manholes captured the intended population at Facility A, F, and G. First, a fluorescent tracer dye (Bright Dyes® FLT yellow/green tablet; Kingscote Chemicals, Miamisburg, OH) was flushed from ≤ 2 toilets inside the facility at different floors and wings, and a stopwatch was used to monitor the time taken to visualize the dye. Time data from the visual tracer was used to establish collection timeframes for the biological tracer study (Table 2). The biological tracer consisted of a heat-inactivated (60±2°C for 40 min) Inforce® 3 Respiratory Vaccine lyophilized calf vaccine (Parsippany, New Jersey; Zoetis) at an eluted final concentration of $10^6$ to $10^{10}$ copies per 50 mL buffer, where bovine respiratory syncytial virus (BRSV) was the target biological tracer. A virus surrogate was chosen out of convenience due to an on-going respiratory wastewater surveillance project. The biological tracer study (n=3 per facility) collected wastewater starting at time $t_0$ and time points were adjusted between study runs to capture resolution in the early peak and signal dissipation (Table 2). To detect the biological tracer, wastewater samples were concentrated and analyzed using ddPCR (Santiago et al. 2025).

Strainer assessment

Three strainers compatible for use with the Teledyne ISCO Avalanche Autosampler (Lincoln, NE) were evaluated at Facility A with the aim of consistent collection of the desired wastewater sample volume (Figure 1). The Standard Weighted Polypropylene Strainer ("Standard", n=9; Lincoln, NE; Teledyne ISCO), Low Flow Stainless Steel Strainer ("Low Flow", n=7; Lincoln, NE; Teledyne ISCO), and a custom fabricated strainer ("Custom", n=9; Atlanta, GA; CDC) were deployed, and 'expected' and 'collected' sample volumes were recorded. See Figure 1 for a strainer visuals and design differences, as well as and the Custom strainer technical drawing in Supplemental Materials 1 (S1). The 'expected' volumes were based upon entered program settings, while 'collected' volumes were measured in the field with graduated sample containers. The sample sizes per strainer were not uniform due to (1) evaluation of one strainer at a time, and a strainer design being removed from evaluation when either (2) clogging continuously disrupted sampling, and/or (3) the 'expected' versus 'collected' were continuously different. Statistical analysis and figure creation used GraphPad version 10.2.0 (Boston, MA), where the absolute values of the 'expected' and 'collected' differences were used to determine the significance between the groups with Kruskal-Wallis and Dunn's post-hoc tests ($p<0.05$).

**B. Feasibility – Individual state results**

Pilot Survey

In the GA (Facilities A to G) pilot survey, all seven participating facilities had a manhole identified for potential sampling (wastewater access availability) and while one facility had an unsafe wastewater collection site, all were amenable to the project and open to communication avenues (i.e., handouts to residents/patients to explain project; Table 3). WWS was deemed feasible at 4 (57.1%) of the 7 GA SNFs.

Expanded Pilot Survey

The expanded pilot survey results regarding general wastewater access, wastewater access location, and facility administration aspects, as well as feasibility concerns, are provided in Table 4, where footnotes provide specifics of access point, safety concerns, and extenuating circumstances that would make WWS not feasible. See S3 for the full expanded survey questions, as only highlighted results are included in the text and Table 4.

*Illinois (IL; Facilities H to L)*: Four of the five LTACHs had external physical access to wastewater (three via a single manhole; one via a combination of manhole and wastewater lift station) that captured all the facilities' flow. These four LTACHs had a manhole location with adequate space for setting up equipment, and one also had an outlet nearby (<20 ft) to power an autosampler. The one facility (Facility J) with no manhole located on the property was not included in any other components of the survey. WWS was deemed feasible at 4 (80.0%) of 5 IL LTACHs.

*Utah (UT; Facility M)*: The RH had external physical access to wastewater via a manhole that captured all the facility wastewater effluent. There was inadequate space to set-up equipment by the manhole due to being located by dumpsters at a loading dock, nor an outlet nearby to power an autosampler. However, there was adequate space within the manhole to deploy an autosampler. WWS was deemed feasible at the UT RH.

*Texas (TX; Facility N)*: The vSNF had external physical access to wastewater via a manhole that captured all the facility wastewater effluent. There was not adequate space to set-up equipment by the manhole, which was in a parking lot behind the facility, adjacent to an access road; and no proximal outlet to power an autosampler was available. However, there was adequate space within the manhole to deploy an autosampler. WWS was deemed feasible at the TX vSNF.

*Arizona (Facilities O and P)*: Both facilities (mixed-use and hospital) had external physical access to wastewater via a manhole that captured all the facility wastewater effluent. The location of the manhole at both facilities had adequate space for setting up equipment, but there were no outlets nearby for either location to power an autosampler. One of the facilities had pedestrian traffic near the manhole, thus a notable safety concern during sampler deployment/retrieval and the other was located in a parking lot within an accessible parking spot requiring planning to ensure the parking space was empty. WWS feasibility was positively determined at both facilities (2 of 2, 100%).

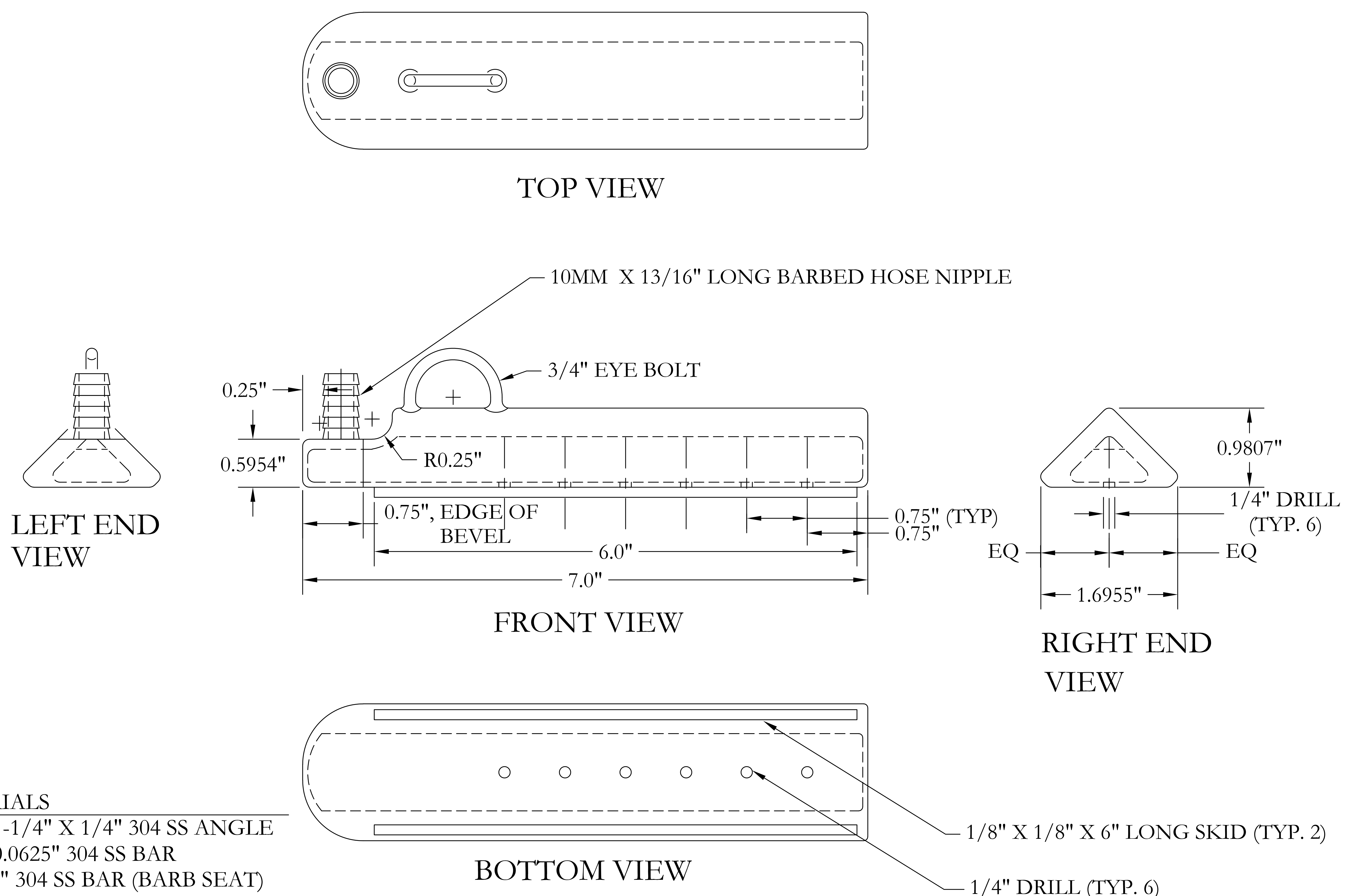

BILL OF MATERIALS

| | |
|---|---|
| 7" | 1-1/4" X 1-1/4" X 1/4" 304 SS ANGLE |
| 7" | 1-1/2" X 0.0625" 304 SS BAR |
| 3" | 1" X 0.125" 304 SS BAR (BARB SEAT) |
| 1-1/2" | 1" X 0.0625" 304 SS BAR (RIGHT END) |
| 2-1/2" | 3/4" X 0.0625 304 SS BAR (NOSE) |
| 13/16" X 10 MM 304 SS BARBED NIPPLE | |
| 3/4" 304 SS U-BOLT | |
| 6" | 1/8" X 1/8" 304 SS BAR |

| REV | DATE | BY |
|---|---|---|

SEAL

U.S. DEPARTMENT OF HEALTH & HUMAN SERVICES
PUBLIC HEALTH SERVICE
CENTERS FOR DISEASE CONTROL AND PREVENTION

SIPHON

DESCRIPTION

PROJECT #: N/A
DATE: 2/13/2023
PROJ. OFFICER: N/A
DRAWN BY: LYY9
CHECKED BY: DSW4

DRAWING #: 1 OF: 1

DWG-001

CONTROLLED
UNCLASSIFIED INFORMATION (CUI)
UNAUTHORIZED DISCLOSURE IS STRICTLY PROHIBITED.
ENHANCED SAFEGUARDING AND DISPOSAL MEASURES ARE REQUIRED.

### Healthcare wastewater expanded pilot survey

**Purpose:** To assess wastewater access at healthcare facilities, particularly skilled nursing facilities and long-term acute care hospitals. These data will be used to understand the feasibility of a national wastewater surveillance program at healthcare facilities. This survey includes an assessment of wastewater access points, the facility's willingness to participate in a wastewater surveillance program, and the safety and feasibility of sampling wastewater at facilities.

| **Section 1: Administrative** | |
|---|---|
| **Questions (8)** | **Answer options** |
| 1. Year facility was built (four-digit year) | ________ (four-digit year)<br>☐ unknown |
| 2. Number of floors in the facility (number) | ________ floors |
| 3. Number of licensed beds or capacity (number) | ________ licensed beds or capacity |
| 4. Census or average occupancy (number) | ________ patients or residents |
| 5. Is the facility amenable to wastewater sampling equipment being set up long-term (i.e., greater than 6 months)? (choose one) | Y☐ / N ☐ / Unsure ☐ |
| 6. Is the facility open to an informational sign on the wastewater sampling equipment and a flier to provide to the public if they have questions? (choose one) | Y☐ / N ☐ / Unsure ☐ |
| 7. Are staff available daily if issues arise?<br>a. Grounds (choose one)<br>b. Plumbing (choose one)<br>c. Engineering (choose one) | a. Y☐ / N ☐ / Unsure ☐<br>b. Y☐ / N ☐ / Unsure ☐<br>c. Y☐ / N ☐ / Unsure ☐ |
| 8. Preferred facility point of contact | Name (text):<br>Phone number:<br>Email address: |

*Source: Coulliette-Salmond et al. JID*

### Healthcare wastewater expanded pilot survey

| Section 2: Willingness to participate | |
|---|---|
| **Questions (4)** | **Answer options** |
| 9. How likely would your facility be to participate in a wastewater monitoring program if it could provide useful information about patient/resident antibiotic resistance burden for infection control? (choose one) | On a scale of 1 (Very likely) to 5 (Very unlikely)<br>1 ☐ Very likely<br>2 ☐ Likely<br>3 ☐ Neutral<br>4 ☐ Unlikely<br>5 ☐ Very unlikely |
| 10. What would make your facility more likely to participate in a wastewater monitoring program? (text) | Describe (text): |
| 11. What are potential barriers to participating in a wastewater monitoring program at your facility? | Describe (text): |
| 12. If applicable, what is the likelihood that your corporate office would support your facility's participation in a wastewater monitoring program? (choose one) | On a scale of 1 (Very likely) to 5 (Very unlikely)<br>1 ☐ Very likely<br>2 ☐ Likely<br>3 ☐ Neutral<br>4 ☐ Unlikely<br>5 ☐ Very unlikely<br><br>☐ Not applicable |

*Source: Coulliette-Salmond et al. JID*

### Healthcare wastewater expanded pilot survey

| Section 3: Wastewater access | |
|---|---|
| **Questions (6)** | **Answer options** |
| 13. Is there at least one external physical access point to the wastewater stream that captures all the facility flow (e.g., sewer manhole on the property)? (choose one) | Y☐ / N ☐ / Unsure ☐<br>If yes, describe (text): |
| 14. Where is/are the external wastewater access point(s) located? (choose all that apply) | ☐ In your parking lot, away from traffic<br>☐ In your parking lot, but where there is traffic<br>☐ In the street, but not in the lanes of traffic<br>☐ In the street and in the lanes of traffic<br>☐ In an area designated for emergency vehicles, deliveries, and transportation services<br>☐ On the landscaped grounds surrounding/next to your building<br>☐ On the sidewalk or in the space between the sidewalk and the street<br>☐ In an area with foot traffic (e.g., entrance, visitor walkway, resident patio)?<br>☐ Unsure<br>☐ Other<br><br>Describe the area (text): |

*Source: Coulliette-Salmond et al. JID*

### Healthcare wastewater expanded pilot survey

| | |
|---|---|
| 15. Approximately how far from the building is/are the wastewater access point(s)? (choose all that apply) | ☐ <20 ft<br>☐ 20-50 ft<br>☐ 50 – 100 ft<br>☐ >100 ft<br>☐ Don't know the location of the manhole(s) |
| 16. Manhole cover size (diameter) and other characteristics:<br>a. Energy source/outlet available? (choose one):<br>b. Description of manhole (e.g., external influences, low point where flooding occurs, near the entrance or parking lot) (text): | a. Y ☐ / N ☐ / Unsure ☐<br>b. Describe (text): |
| 17. Is there sufficient space for setting up the following equipment and for 1 to 3 project staff to work near an appropriate wastewater access point?<br>a. Autosampler (choose one)<br>b. Passive (choose one)<br>c. Other (choose one) | a. Y☐ / N ☐ / Unsure ☐<br>b. Y☐ / N ☐ / Unsure ☐<br>c. Y☐ / N ☐ / Unsure ☐<br>If yes to 2c, describe (text): |
| 18. Assess the sampling location for any feasibility concerns and describe (text): | Describe (text): |

*Source: Coulliette-Salmond et al. JID*

### Healthcare wastewater expanded pilot survey

| Section 4: Grease traps/drainage lines | |
|---|---|
| **Questions (7)** | **Answer options** |
| 19. Does your facility have an external or outside grease trap? (choose one) | Y☐ / N ☐ / Unsure ☐ |
| 20. Does your facility have a wastewater drainage line (lateral) to the sewer that is separate from the grease trap? (choose one) | Y☐ / N ☐ / Unsure ☐ |
| 21. Is the sample location intermingled with the flow to or from the grease trap? (choose one) | Y☐ / N ☐ / Unsure ☐ |
| 22. How many wastewater drainage lines (laterals) other than the grease trap leave the building? (number) | ____________(number) |
| *The questions below are for the first drainage line. (Repeat the following questions for the number of drainage lines)* | |
| 23. Is there a clean out that can be used to access the drainage pipe outside the facility that is associated with the manhole? (choose one) | Y☐ / N ☐ / Unsure ☐ |
| 24. Approximate diameter of the clean out (number)<br>a. Diameter (number)<br>b. Units of diameter (choose one) | a. ________ (number)<br>b. Inches ☐ / Centimeters ☐ |
| 25. Is the clean out horizontal or vertical? (choose one) | ☐ Horizontal<br>☐ Vertical<br>☐ Unsure |
| *Repeat the section above as needed based on number of drainage lines.* | |

*Source: Coulliette-Salmond et al. JID*

### Healthcare wastewater expanded pilot survey

| Section 5: Plumbing questions | |
|---|---|
| **Questions (3)** | **Answer options** |
| 26. Potable water meter location (text) | Describe (text): |
| 27. Is laundry done onsite? (choose one) | Y☐ / N ☐ / Unsure ☐ |
| 28. Does the facility plumbing system have any bleach (chlorine) injectors installed with the laundry or to treat wastewater before it enters the county sewer system? (choose one) | Y☐ / N ☐ / Unsure ☐<br>If yes, specify area where installed (text): |

**Definitions**

Bleach injector: Also known as a chlorine injector; the method of adding or mixing chlorine solution into wastewater using an injector pump

Clean out: An access point in the main sewer line for the purposes of cleaning and unclogging the sewer line, installed outside the facility

Grease trap: Trap or waste pipe designed to prevent most greases and solids from entering a wastewater system

Wastewater: Water that has come in contact with human body wastes such as from toilets or other receptacles, as well as water from showers, sinks, drains, etc. This water typically contains pollutants and pathogens that would negatively impact receiving waters unless treated.

Wastewater monitoring: A consistent collection of wastewater and testing of the wastewater for desired targets (e.g., virus, bacteria, genes) over a defined period

Wastewater sampling equipment: Refers to equipment used to collect wastewater and measure characteristics of wastewater; examples include an autosampler and components (e.g., tubing, pump), flow meter, instruments to measure temperature and pH

*Source: Coulliette-Salmond et al. JID*
